# Selective tweeting of COVID-19 articles: Does title or abstract positivity influence dissemination?

**DOI:** 10.1101/2021.06.22.21259354

**Authors:** Nicholas Fabiano, Zachary Hallgrimson, Stanley Wong, Jean-Paul Salameh, Sakib Kazi, Rudy R Unni, Lee Treanor, Robert Frank, Ross Prager, Matthew DF McInnes

## Abstract

**Background:** Previous research has shown that articles may be cited more frequently on the basis of title or abstract positivity. Whether a similar selective sharing practice exists on Twitter is not well understood. The objective of this study was to assess if COVID-19 articles with positive titles or abstracts were tweeted more frequently than those with non-positive titles or abstracts.

**Methods:** COVID-19 related articles published between January 1^st^ and April 14^th^, 2020 were extracted from the LitCovid database and all articles were screened for eligibility. Titles and abstracts were classified using a list of positive and negative words from a previous study. A negative binomial regression analysis controlling for confounding variables (2018 impact factor, open access status, continent of the corresponding author, and topic) was performed to obtain regression coefficients, with the p values obtained by likelihood ratio testing.

**Results:** A total of 3752 COVID-19 articles were included. Of the included studies, 44 titles and 112 abstracts were positive; 1 title and 7 abstracts were negative; and 3707 titles and 627 abstracts were neutral. Articles with positive titles had a lower tweet rate relative to articles with non-positive titles, with a regression coefficient of -1.10 (P < .001), while the positivity of the abstract did not impact tweet rate (P = .2218).

**Conclusion:** COVID-19 articles with non-positive titles are preferentially tweeted, while abstract positivity does not influence tweet rate.

## Introduction

Coronavirus disease 2019 (COVID-19) was first detected in Wuhan, China in December 2019, and its rapid spread has resulted in a global pandemic (1,2). The global impact of the COVID-19 pandemic has elicited a response from scientists and researchers worldwide with over 5000 COVID-19 related articles listed on PubMed as of April 18^th^, 2020 (3,4). This expanding volume of research has necessitated streamlined methods to disseminate information to frontline clinicians in a timely manner. As a result, scientists and clinicians have increasingly turned to social media platforms such as Twitter to share, discuss, and respond to peer reviewed studies, in some cases prior to the peer review process (5).

Social media has become integrated within the research workflow with applications throughout the continuum of research reporting, from the identification of opportunities to dissemination of findings (6,7). Traditionally, citation-based metrics have been employed in order to quantify and track the dissemination of research findings; however, these metrics neglect the dissemination of a study online through social media. Altmetric is an online tool which tracks social media sharing of articles from a variety of sources such as Twitter, Facebook, and Reddit (8,9). Altmetric has previously been used to explore how variables such as study discipline, number of authors, and country of origin are associated with an article’s dissemination online (10,11). Studies have shown a lack of correlation between Twitter-based altmetrics and citation-based metrics, suggesting that altmetrics reflect a distinct yet complementary source of information to traditional citation-based metrics (12,13). Thus, the dissemination of a particular study can be more holistically assessed through the incorporation of altmetrics (14,15). Particularly, citation-based metrics are subject to a time-lag during times of rapid research output, such as the COVID-19 pandemic, due to the publication process. Altmetrics are not subject to this time-lag, which gives them potential utility to track the rapid dissemination of research.

Understanding the determinants of research dissemination is essential, as various study characteristics may lead to the preferential sharing of certain findings. For example, a particular study may be cited or shared online more frequently on the basis of positive findings. These selective citation practices are well documented in medical literature and are particularly problematic since studies with positive results may be overrepresented, potentially influencing the perceived effectiveness of a particular intervention and leading to a systematic deviation from the truth; known as citation bias (16–18). Particularly, it has been found that studies with positive titles or conclusions, or higher accuracy estimates are cited more frequently in imaging diagnostic accuracy literature (19,20). It is not well understood whether an analogous form of selective tweeting exists in COVID-19 literature. The objective of this study is to assess for the presence of selective tweeting in COVID-19 literature by evaluating whether articles with positive titles or abstracts are tweeted more frequently than those with negative or neutral titles or abstracts.

## Methods

Research ethics board approval for this type of research is waived at the University of Ottawa. The pre-specified study protocol is available at the following link https://osf.io/5wvjq/

### Search strategy and selection

LitCovid (3), a curated repository of COVID-19 research that is updated on a daily basis, was searched on April 11^th^, 2020 for all publications included in the database to date.

Two investigators (NF [second year medical student] and ZH [third year medical student]) independently screened each of the retrieved articles for potential relevance in duplicate and disagreements were resolved by consensus. Articles were excluded if they were not on the topic of COVID-19, the title was not in English, or if they were retracted.

### Data collection

Four investigators (ZH, SW [second year medical student], SK [second year medical student] and LT [third year medical student]) extracted the article title, abstract, authorship, publication date, continent of corresponding author (Asia, Africa, North America, South America, or Australia), and topic (General Info, Mechanism, Diagnosis, Transmission, Treatment, Prevention, Epidemic Forecasting, and Case Report) from LitCovid. Two investigators (NF and SW) extracted the 2018 journal impact factor and open access status (open access or subscription-based) from Clarivate Analytics Journal Citation Reports. One investigator (ZH) used Altmetric to derive the number of tweets made about an article (on April 18^th^, 2020). Article DOIs were used to query the Altmetric Details Page API, version v1. If DOIs were not available, the article title was used to search Altmetric Explorer. All extractions were done independently, in single. Missing information was recorded as “not reported”. Default values were assigned in cases where the complete publication date was not available: in cases where the day of publication was unavailable, the first of the month was assigned as default, and in cases where only the year of publication was available, March 1^st^ was assigned as default. For journals not listed on Clarivate Analytics Journal Citation Reports an impact factor of 0 was assigned. For studies with multiple topics or continents listed, the first listed was extracted.

Five investigators analyzed the full text of the articles independently and in duplicate (NF [50%], ZH [50%]; SW [70%], SK [15%], and LT [15%]) to determine type of article (original research or not original research), and disagreements were resolved by consensus. Articles whose full text was not available through institutional subscription or whose abstract was not in English were classified as “unspecified”.

### Assessment of title and abstract positivity

Sentiment criteria was derived and modified from a previous study (21). Positive, and negative categories contained 25 words each and all other words were deemed to be neutral, as shown in Figure 1. Python v3.7.4 was used to search included article titles or abstracts for words in sentiment criteria. All words were assigned as +1 (positive), -1 (negative), or 0 (neutral) based on the previously described categories. The positive word ‘novel’ was deemed neutral in all cases referring directly to the name of the virus itself. If the sum was greater than 0, less than 0, or equal to 0, the abstract or title was deemed to be positive, negative, or neutral, respectively. If no abstract was present, a default value of 0 (neutral) was assigned.

**Figure 1.**
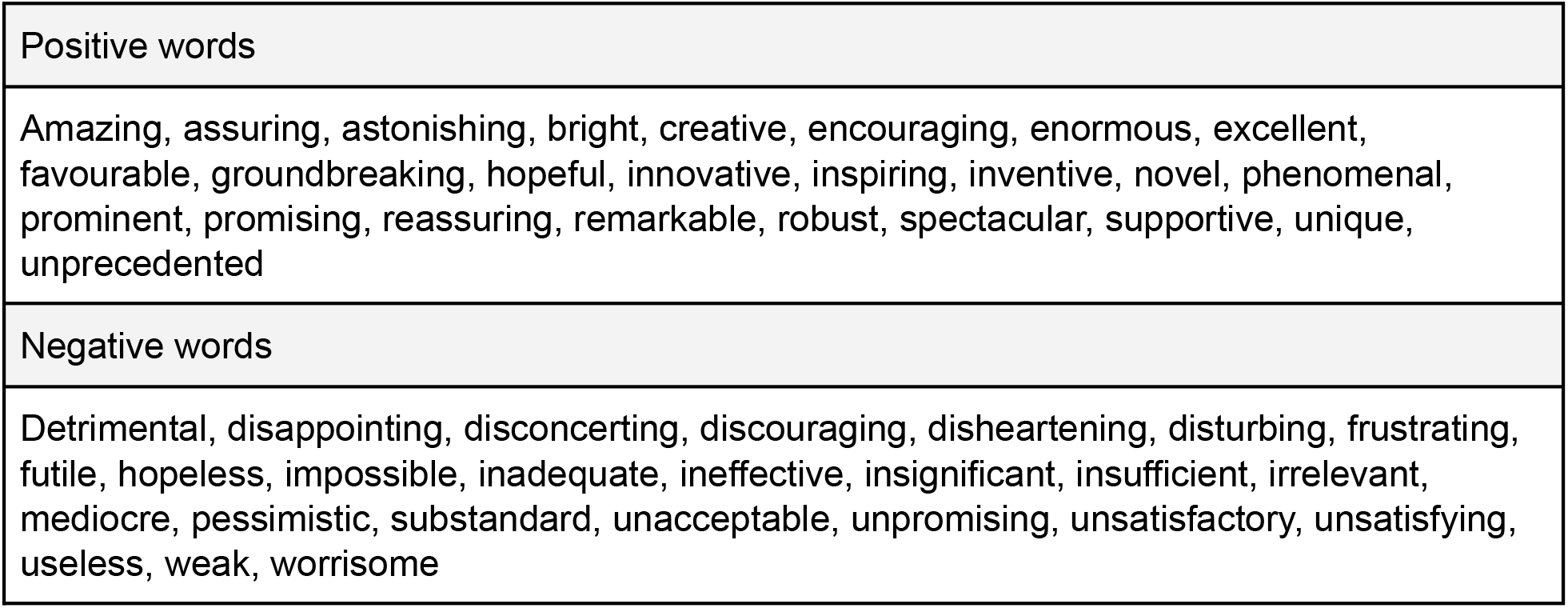
Sentiment Criteria.

### Data analysis

A negative binomial regression analysis was conducted to evaluate the association between title positivity and tweet rate, controlling for the following confounding variables: open access status, continent of corresponding author, topic and 2018 journal impact factor. Inclusion of variables into the final model was performed using stepwise selection based on the Akaike information criterion (AIC) in R (MASS package) (22–24). A subgroup analysis was then conducted for original and non-original research articles. For the original research articles, another negative binomial regression analysis was used to evaluate the relationship between abstract positivity and tweet rate, controlling for the same variables as defined previously. For the non-original articles, a negative binomial regression analysis was conducted looking at tweet rate and controlling for the same confounding variables. Articles classified as “unspecified” were excluded from the subgroup analysis

Regression coefficients were calculated for each outcome variable to evaluate the association between a specific variable and tweet rate. Positive and negative regression coefficients represented positive and negative associations between a given variable and tweet rate, where the magnitude of the regression coefficient represented the strength of association. Predictive Mean Matching imputation was performed for variables with more than 10% missingness proportion. The imputation was performed with the MICE package in R (version 3.6.3, R Foundation for Statistical Computing) (25–28). The p-values for the association of the variables of interest with tweet rate were obtained using R software by means of likelihood ratio testing with a chi-squared mode, where a value of 0.05 was used as the threshold for significance.

## Results

### Study characteristics

Of the 3761 articles extracted from LitCovid, 3752 articles remained after application of the exclusion criteria, as outlined in Figure 2. Included study characteristics are presented in Table 1 and 2. Twitter summary characteristics based on title positivity is presented in Table 3. Publication dates ranged from January 1, 2020 to April 14, 2020. 434 (11.51%) articles were assigned a default day and 102 (2.71%) were assigned a default day and month. 889 separate journals were represented in the study population with the most common journal being BMJ with 200 articles (5.45%). 797 articles (21.19%) had multiple topics and were assigned the first topic listed.

**Table 1.**
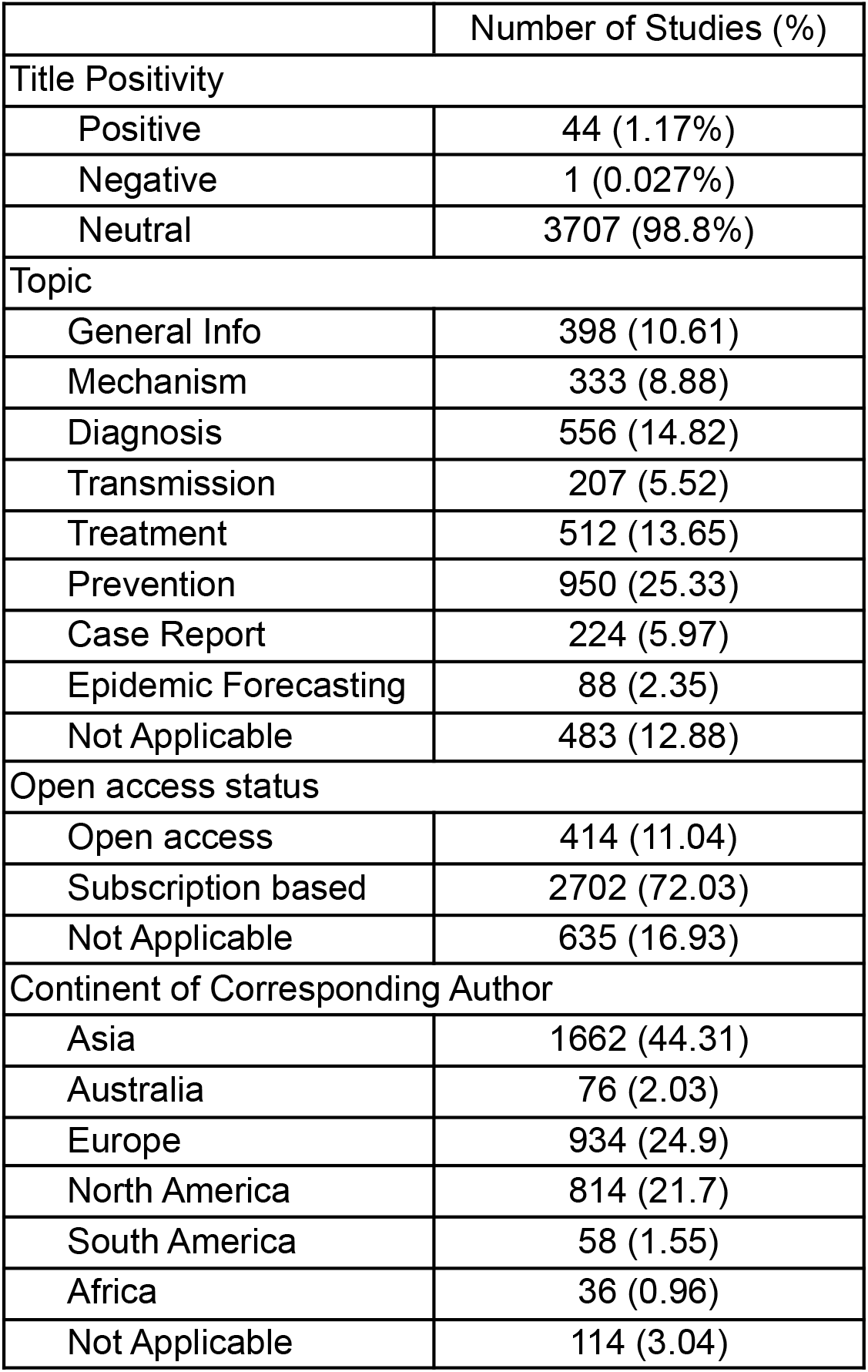
Summary characteristics for all 3752 included studies.

|  | Number of Studies (%) |
| --- | --- |
| Title Positivity |  |
| Positive | 44 (1.17%) |
| Negative | 1 (0.027%) |
| Neutral | 3707 (98.8%) |
| Topic |  |
| General Info | 398 (10.61) |
| Mechanism | 333 (8.88) |
| Diagnosis | 556 (14.82) |
| Transmission | 207 (5.52) |
| Treatment | 512 (13.65) |
| Prevention | 950 (25.33) |
| Case Report | 224 (5.97) |
| Epidemic Forecasting | 88 (2.35) |
| Not Applicable | 483 (12.88) |
| Open access status |  |
| Open access | 414 (11.04) |
| Subscription based | 2702 (72.03) |
| Not Applicable | 635 (16.93) |
| Continent of Corresponding Author |  |
| Asia | 1662 (44.31) |
| Australia | 76 (2.03) |
| Europe | 934 (24.9) |
| North America | 814 (21.7) |
| South America | 58 (1.55) |
| Africa | 36 (0.96) |
| Not Applicable | 114 (3.04) |

**Table 2.** Twitter and impact factor summary characteristics for included studies.

|  | Total tweets | Tweet rate<br>(tweets/day) | Impact factor |
| --- | --- | --- | --- |
| Mean | 309.72 | 12.54 | 11.13 |
| Median | 16 | 0.65 | 3.704 |
| Range - Max | 39856 | 1811.64 | 70.76 |
| Range - Min | 0 | 0 | 0 |
| Standard deviation | 1454.91 | 62.3 | 16.47 |
| Variance | 2116759.4 | 3880.95 | 271.11 |

**Table 3.** Twitter summary characteristics based on title positivity for all included studies.

|  | Positive |  | Neutral |  | Negative |  |
| --- | --- | --- | --- | --- | --- | --- |
|  | Total tweets | Tweet rate (tweets/day) | Total tweets | Tweet rate (tweets/day) | Total tweets | Tweet rate (tweets/day) |
| Mean | 44.95 | 2.42 | 313.26 | 12.67 | 71 | 2.51 |
| Median | 7.5 | 0.26 | 17 | 0.66 | 4.5 | 0.26 |
| Range - Max | 620 | 38.75 | 39856 | 1811.64 | 364 | 11.74 |
| Range - Min | 0 | 0 | 0 | 0 | 0 | 0 |
| Standard deviation | 103.47 | 6.80 | 1464.33 | 62.70 | 144.91 | 4.64 |
| Variance | 10706.60 | 46.24 | 2144272.9 | 3931.43 | 20999.2 | 21.56 |

**Figure 2.**
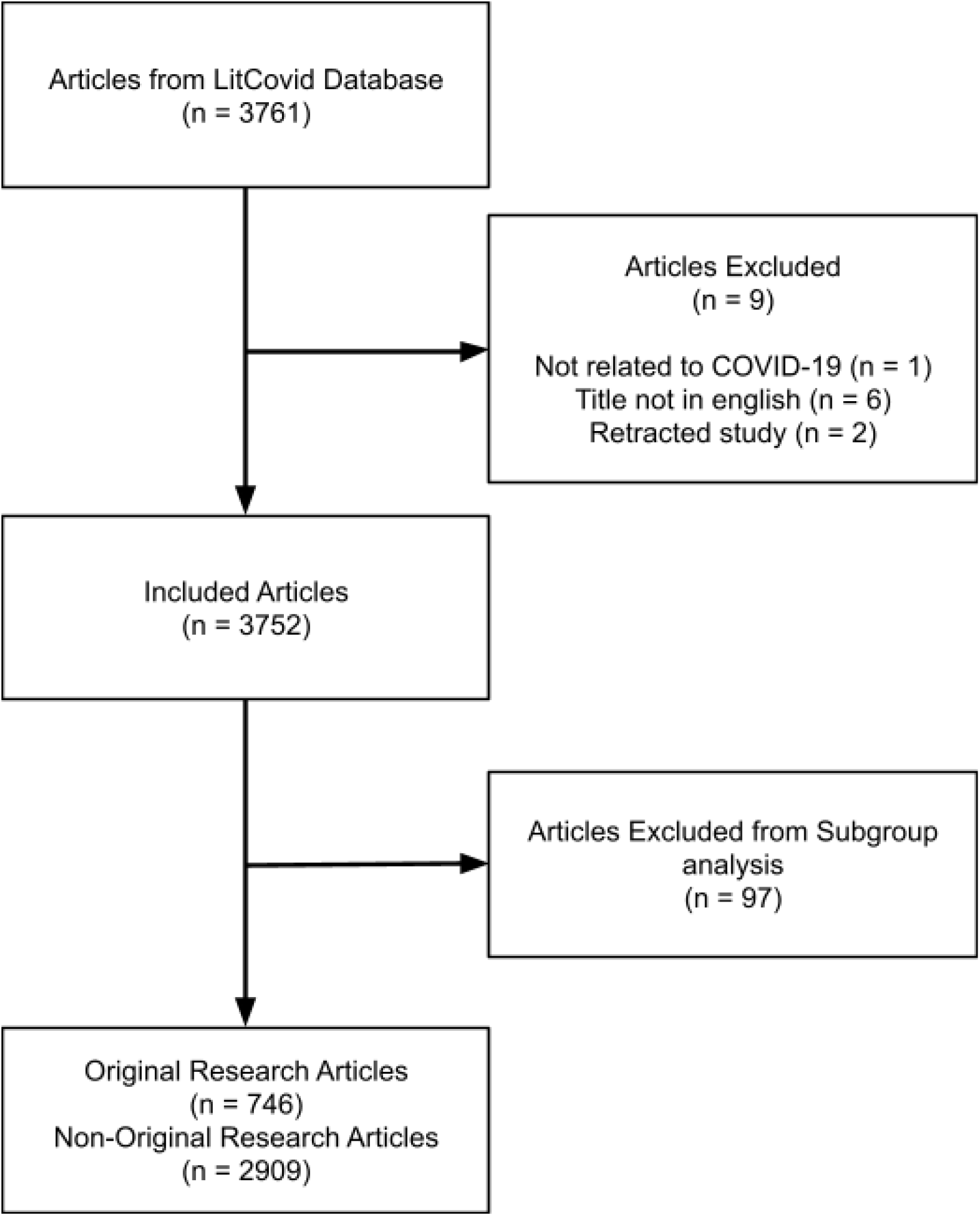
Flow diagram outlining article selection and exclusions.

Within the original research articles, there were 13 articles with positive titles, 0 articles with negative titles and 733 articles with neutral titles. Further, there were 112 positive, 7 negative and 627 neutral abstracts. Twitter summary characteristics based on abstract positivity are presented in Table 4. For the non-original articles, there were 31 positive, 6 negative and 3902 neutral titles.

**Table 4.** Twitter summary characteristics based on abstract positivity for all original research studies.

|  | Positive |  | Neutral |  | Negative |  |
| --- | --- | --- | --- | --- | --- | --- |
|  | Total tweets | Tweet rate (tweets/day) | Total tweets | Tweet rate (tweets/day) | Total tweets | Tweet rate (tweets/day) |
| Mean | 535.26 | 21.37 | 554.04 | 22.96 | 124.57 | 6.37 |
| Median | 44.5 | 1.16 | 19 | 0.72 | 7 | 0.27 |
| Range - Max | 14750 | 543.5 | 39856 | 1811.64 | 740 | 41.11 |
| Range - Min | 0 | 0 | 0 | 0 | 0 | 0 |
| Standard deviation | 1962.8 | 67.83 | 2437.48 | 107.59 | 274.01 | 15.33 |
| Variance | 3852591.22 | 4600.84 | 5941313.11 | 11574.8 | 75081.95 | 235.09 |

### Association of title or abstract positivity with tweet rate

A positive relationship was seen for all included studies between tweet rate and impact factor, open access status and the topics general info, prevention, transmission and treatment. For all included studies a negative relationship was seen between tweet rate and title positivity. For the original articles, a positive relationship was seen between tweet rate and impact factor, and the continent Europe with a negative relationship seen between tweet rate and title positivity, open access status and the continent Africa. For non-original research articles, a positive relationship can be seen between tweet rate and impact factor, open access status, and the continent Europe, with a negative relationship seen between tweet rate and title positivity. From this it is seen that for the total dataset and non-original research open access articles are associated with lower tweet rates, while for original research the open access articles are associated with a higher tweet rate. The regression coefficients can be seen in Table 5.

**Table 5.** Regression coefficients of several variables, including title positivity, with tweet rate as determined by negative binomial regression analysis. Variables without an associated regression coefficient were excluded based on stepwise analysis and labelled as “NA”.

|  | Total dataset |  | Original research articles |  | Non-original research articles |  |
| --- | --- | --- | --- | --- | --- | --- |
|  | Regression coefficient (SE) | P-value | Regression coefficient (SE) | P-value | Regression coefficient (SE) | P-value |
| Intercept | -2.74 (0.14) | <.001 | -1.74 (0.13) | <.001 | -2.86 (0.13) | <.001 |
| 2018 Impact Factor | 3.09 (.19) | <.001 | 3.57 (.36) | <.001 | 2.59 (.21) | <.001 |
| Title Positivity |  |  |  |  |  |  |
| Positive | -1.10(.33) | <.001 | -1.65(.62) | .01 | -1.14(.382) | .003 |
| Non-Positive | REF | NA | REF | NA | REF | NA |
| Open Access Status |  |  |  |  |  |  |
| Subscription based | 2.44 (0.07) | <.001 | -1.12 (.40) | <.001 | 2.46 (.08) | <.001 |
| Open access | REF | NA | REF | NA | REF | NA |
| Continent of corresponding author |  |  |  |  |  |  |
| Australia | NA | NA | -2.71(.94) | .004 | .46(.25) | .05 |
| Europe | NA | NA | 1.10 (.23) | <.001 | .46 (.09) | <.001 |
| Africa | NA | NA | -4.60 (1.14) | <.001 | .035 (.35) | .92 |
| North America | NA | NA | .59 (.22) | .01 | .21(.09) | .03 |
| South America | NA | NA | -0.71 (0.69) | 0.31 | 0.26 (0.29) | 0.39 |
| Asia | NA | NA | REF | NA | REF | NA |
| Topic |  |  |  |  |  |  |
| Diagnosis | .27 (.15) | .07 | NA | NA | .28 (.16) | .09 |
| Epidemic Forecasting | -.09 (.20) | .67 | NA | NA | .75 (.30) | .01 |
| General Info | .67 (.16) | <.001 | NA | NA | .02 (0.16) | .89 |
| Mechanism | -.02 (.16) | .92 | NA | NA | -.38(.18) | .04 |
| Prevention | .54 (.14) | <.001 | NA | NA | .21 (.14) | .14 |
| Transmission | 1.30 (.18) | <.001 | NA | NA | -.01 (.20) | .97 |
| Treatment | .68 (.15) | <.001 | NA | NA | -.35 (.16) | .03 |
| Case Report | REF | NA | NA | NA | REF | NA |

For the original research article abstracts, a positive relationship was seen between tweet rate and impact factor with a negative relationship seen between tweet rate and open access status. The variables of the author’s corresponding continent and topic were eliminated based on stepwise analysis. The regression coefficients can be seen in Table 6.

**Table 6.** Regression coefficients for original article abstracts looking at 2018 impact factor, abstract positivity, and open access status compared with tweet rate as determined by negative binomial regression analysis.

|  | Regression coefficient (SE) | P-value |
| --- | --- | --- |
| Intercept | -1.42 (0.13) | <.001 |
| 2018 Impact Factor | 3.79 (.36) | <.001 |
| Abstract Positivity |  |  |
| Positive | -.04 (.22) | .87 |
| Non-positive | REF | NA |
| Open Access Status |  |  |
| Subscription based | -1.44 (.40) | <.001 |
| Open access | REF | NA |

## Discussion

### Principal findings

The objective of this study was to assess for the presence of selective tweeting in COVID-19 literature by assessing if title or abstract positivity influences the number of times an article is tweeted. A negative association was found between tweet rate and positive titles meaning that studies with positive titles were tweeted less frequently than those with non-positive ones. This association persisted in the sub analyses for original and non-original research articles. For the original research articles, no association was found between abstract positivity and tweet rate. Further, there were substantially fewer negative titles (n = 1) and abstracts (n = 7) than positive titles (n = 44) and abstracts (n = 112). These findings are consistent with the current state of several scientific disciplines which have a low prevalence of negative studies and a high prevalence of positive studies (29,30).

Surprisingly, our findings suggest that COVID-19 articles with positive titles are tweeted to a lesser extent than those with non-positive titles. Previous research has shown that positive titles and abstracts are known drivers of citations, and since tweets and citations are weakly associated, one may infer that positive titles and abstracts would also serve as drivers for tweets (13,19). Despite this, the opposite was observed which may indicate that there are other drivers of tweet rate which dominate such as the 2018 impact factor, open access status, continent of the corresponding author, or topic. Particularly, higher impact factor, articles in subscription based journals, and several topics (General Info, Prevention, Transmission, and Treatment) were found to be positively associated with tweet rate. Further, articles with overly positive titles may be perceived as a form of ‘academic clickbait’, and previous research has found that these strategies negatively influence their perceived quality, causing readers to avoid them (31–33). This effect may be magnified during the COVID-19 pandemic, where the management of such crises relies on the dissemination of credible information.

Interestingly, although a significant association was present between title positivity and tweet rate, there was a lack of significant association between abstract positivity and tweet rate. This finding could indicate that a large sum of readers share articles on the basis of the title without taking the time to read the contents of the article itself, let alone the abstract. This theory is supported by a study which found that 59% of articles shared on Twitter are shared without ever being read. (34). The sharing of articles on Twitter on the basis of title, without reading the contents of the article itself, may allow for the dissemination of poor quality research.

### Limitations

This study is subject to limitations. Firstly, the list of positive and negative words is finite, and thus may not have quantified all sentiment present in the titles and abstracts. A prespecified list of words was employed in order to exclude the subjective bias in classifying the sentiment of words as positive, negative, or neutral. Further, the location or context of the word in the title or abstract was not studied, which could hold significant value. Moreover, self-promotion (e.g. authors tweeting their own article) could influence the number of tweets an article receives, however, the extent of this is difficult to quantify. Also, this study did not determine the sentiment of the tweets, which could provide insightful information. Despite this, it has been demonstrated that 94.8% of tweets linking to scientific papers were neutral in sentiment (35), suggesting that a similar trend was present in this cohort. There was also a loss of precision due to the use of the categorical variable continent instead of the country of the corresponding author’s institution, which was necessary in order to get an appropriate fit of the model. Similarly, although imputation was performed to address the relatively high proportion of missing values, this could have impacted the robustness of the model fit to the data.

## Conclusion

As Twitter becomes increasingly integrated within the research workflow, the selective tweeting of articles with positive titles or abstracts can mislead researchers and clinicians as they seek evidence for the management of COVID-19. This study demonstrated that positive titles are associated with lower tweet rates than non-positive titles, and that abstract positivity does not impact tweet rate. Further research should aim to assess for the presence of selective citation practices in COVID-19 literature.

## Data-Sharing Statement

All articles were extracted from the LitCovid database. Data is available upon request.

## Data Availability

The pre-specified study protocol is available at the following link https://osf.io/5wvjq/

## Declarations

### Ethical approval

Research ethics board approval for this type of research is waived at the University of Ottawa.

### Guarantor

Dr. M McInnes

### Contributorship

Nicholas Fabiano and Zachary Hallgrimson researched literature, conceived the study and participated in data collection, analysis and manuscript creation. Stanley Wong was involved in data collection, analysis and manuscript creation. Jean-Paul Salameh was involved in data analysis. Rudy Unni and Lee Treanor helped in data collection and manuscript editing. Ross Prager participated in study design, data analysis and manuscript editing. Matthew McInnes supervised the project, contributing to study design and manuscript writing and editing. All authors reviewed and edited the manuscript and approved of the final manuscript version.

## Acknowledgements

We would like to thank all who participated in this project.

